# Features and Networks of the Mandible on Computed Tomography

**DOI:** 10.1101/2023.08.04.23293634

**Authors:** Tuan D. Pham, Simon B. Holmes, Mangala Patel, Paul Coulthard

**Affiliations:** Barts and The London School of Medicine and Dentistry, Queen Mary University of London, Turner Street, London E1 2AD, UK

**Keywords:** Mandible, computed tomography, image processing, nonlinear data analysis, network theory, artificial intelligence

## Abstract

The mandible or lower jaw is the largest and hardest bone in the human facial skeleton. Fractures of the mandible are reported to be a common facial trauma in emergency medicine and gaining insights into mandibular morphology in different facial types can be helpful for trauma treatment. Furthermore, features of the mandible play an important role in forensics and anthropology for identifying gender and individuals. Thus, discovering hidden information of the mandible can benefit interdisciplinary research. Here, for the first time, a method of artificial intelligence-based nonlinear dynamics and network analysis are utilized for discovering dissimilar and similar radiographic features of mandibles between male and female subjects. Using a public dataset of ten computed tomography scans of mandibles, the results suggest a difference in the distribution of spatial autocorrelation between genders, uniqueness in network topologies among individuals, and shared values in recurrence quantification.

## 1 Introduction

The mandible known as lower-jaw bone is multi-functional. It constitutes the shaping of the lower face, regulation of mastication, and accountability for the articulation of phonemes. In facial trauma, a severe injury to the mandible can cause serious adversarial impact on the patient. Thus, understanding characteristics of the mandible such as its morphology, physiology, and genetic profile can be helpful for improving mandibular reconstruction. As a result, it can accelerate the healing process and prevent the patient from postsurgical complications [1].

On another scientific aspect, the ability to determine sex from an unknown human bone plays an important role in forensics and anthropology. Because the mandible can withstand the effect of postmortem damage and provide informative evidence about sexual traits, it is commonly used in these fields [2]. Cross-sectional slices of mandibles on cone beam computed tomography (CT) of two young cohorts of 268 males and 386 females were used to discover different morphological characteristics of sexes with nonlinear regression analysis of the mandibular growth trajectories [3]. Insights gained from the study can lead to better abilities for assessing craniofacial abnormalities, identifying age and sex of juvenile remains. An investigation [4] reported that 14 out of 16 radiographic data of the adult mandible could be used for identifying sexual dimorphism, and 15 out of 20 morphometric studies of the dry mandible suggested a relationship between mandibular parameters and sexual dimorphism. Another work found a significant difference in the mandibular angle between males and females [5]. The authors concluded that gonial angle, and antegonial angle and depth can be utilized as measurements of forensic evidence for gender determination.

On a clinical aspect, craniofacial abnormalities are known to be a risk factor associated with obstructive sleep apnea (OSA). However, this association still needs to be validated with physical evidence. A study [6] used CT scans to measure mandibular width and other parameters of the head and neck of a cohort. The study found a correlation between the mandibular width and OSA, and wide mandibles are subject to attention for screening of the risk. Temporomandibular disorders (TMDs) are a group of pathologies that cause pain and dysfunction around the temporomandibular joint and muscles regulating the jaw movement, particularly when chewing. A study aimed to describe the mandibular postural position and mouth opening in healthy subjects and patients with articular/muscular pathology for discovering novel characteristics of TMDs [7], such study has highlighted the importance for extracting useful information from the mandible. On a more dental aspect, a study [8] tried to determine the normal size of the mandible and investigated difference between the dental arch length and total teeth size space. The finding can help prevent the sharp collision of the lower third molars by performing a preventive or therapeutic surgery. Furthermore, the same study highlighted the important role of the mandibular morphology for determining aesthetics and recommending patients for orthognathic and reconstructive surgeries.

Being motivated at discovering novel features of the mandible in radiology, this paper introduces an original study in coupling the theory of spatial statistics and artificial intelligence (AI)-based nonlinear dynamics for discerning subtle dissimilarity and similarity of mandibles on CT scans obtained from male and female subjects. The rest of this paper is organized as follows. Section 2 describes a public CT dataset of mandibles for testing the proof of concept. To make the highly interdisciplinary content of this paper self-contained by providing adequate background, Section 3 outlines technical methods adopted in this study for extracting new mandibular imaging features. Such methods include geostatistics as a branch of spatial statistics, fuzzy recurrence plots and their quantitative measures of nonlinear dynamics. Section 4 shows results of the analysis together with discussion of the finding. Finally, Section 5 concludes the study with emphasis and limitation of the current finding as well as suggested issues for future work.

## 2 Data

The image data used in this study were provided and described in [9], aiming to facilitate serial image studies of the human mandible and assessment of automated segmentation algorithms. The data consist of 10 anonymized CT scans of the human craniomaxillofacial complex originally in DICOM (Digital Imaging and Communications in Medicine) format files. The image slices provide complete physiological mandibles and mandibular bone structures without teeth and with the inclusion of atrophic and non-atrophic mandibular bones.

The 10 CT scans were from 5 male and 5 female subjects out of 45 original CT scans. This selection was to ensure high-quality ground-truth data having clear bone contours and anatomical structures without artefacts. The data are publicly available for downloading from a Figshare repository [10], where CT scans as stacked TIFF (Tag Image File Format) files are provided and used in this study.

Table 1 briefly lists the CT-scan and demographic information of the five female (S1-S5) and five male subjects (S6-S10). Figure 1 shows the montages of 25 CT slices (75th to 100th) of female subject S1 and male subject S6.

**Table 1:**
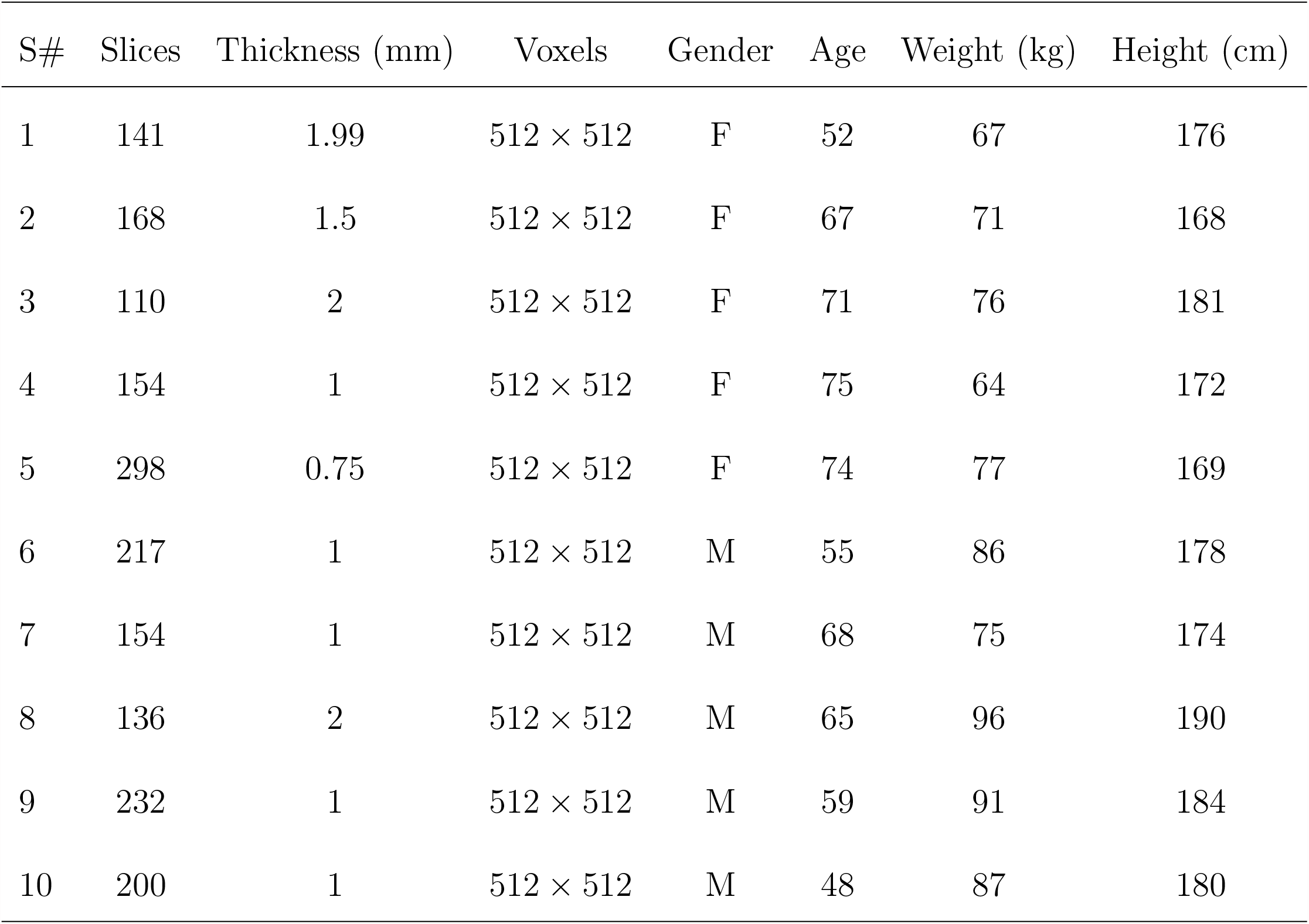
CT scans and subjects’ demographics.

**Figure 1:**
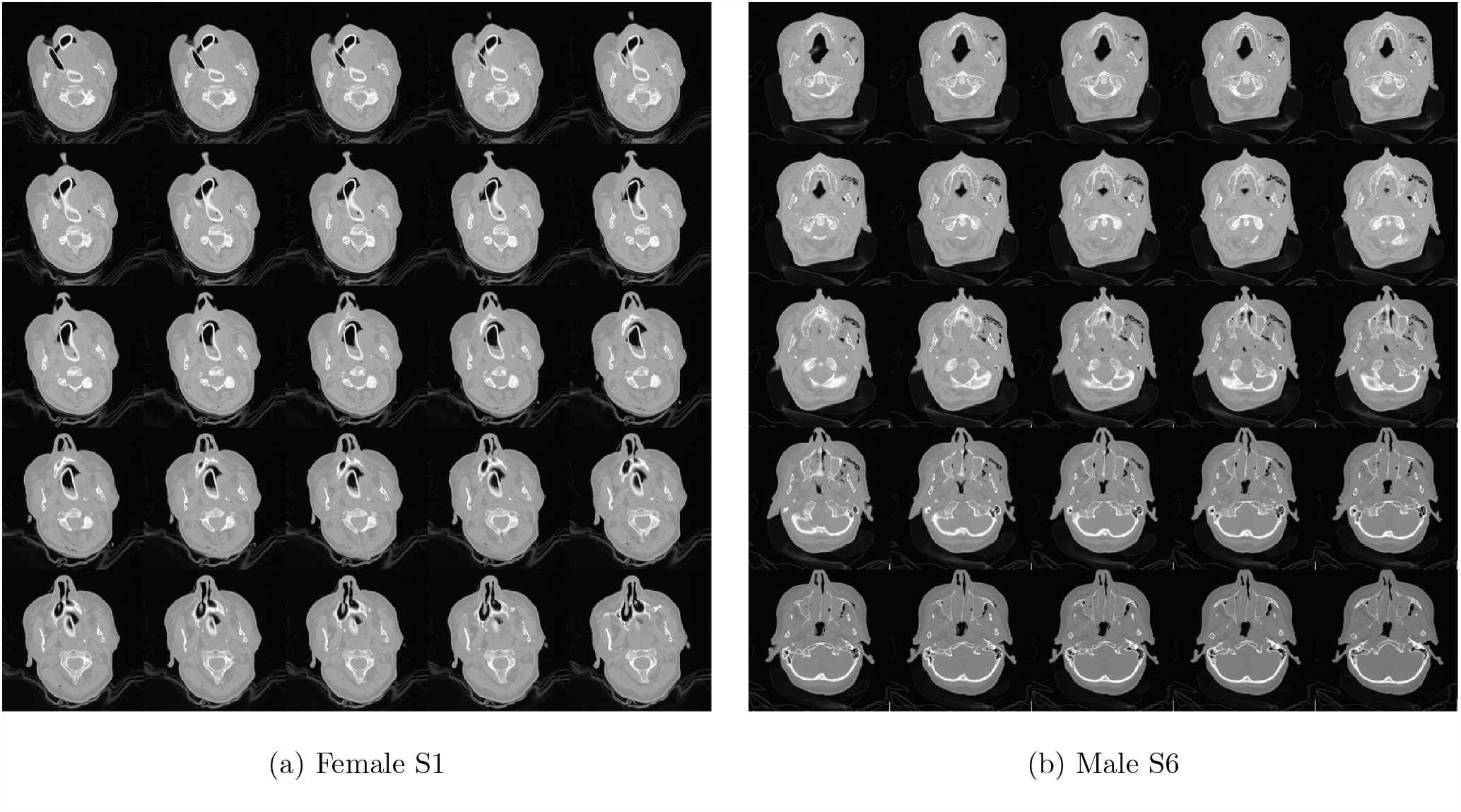
Montages of CT slices obtained from female and male subjects.

## 3 Methods

### 3.1 Geostatistics

As a class of spatial statistics, geostatistics can be applied to quantify variances of variables associated with spatial or spatiotemporal phenomena. It is used in this study to determine spatial variances of image intensities in sequential CT slices of the mandibles. Let *f* (*x, y*) be the intensity value of a pixel on a 2D CT slice *I*(*x, y*). To simplify the mathematical expression, let *f* (*i*) = *f* (*x, y*), where *i* refers to a position in *I, i* = 1, …, *M*, and *M* is the number of pixels in *I*. Geostatistics computes the semi-variogram of variable *f* (*i*) as a function of lag *h*, denoted as *γ* (*h*), as

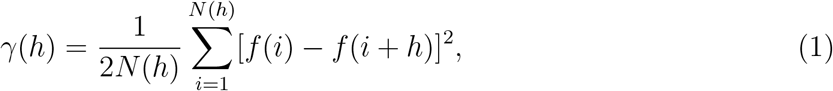

where *N* (*h*) is the number of pairs of *f* (*i*) and *f* (*i* + *h*) that are taken into account in the calculation.

If *γ* (*h*) is computed for each CT slice for a given *h*, then a distribution or time series of *γ* (*h*) for a CT scan be visualized on a plot of *γ* (*h*) or spatial variances against CT slices. Not only series of the CT spatial variance can provide information to discern the difference of image features between male and female mandibles, they can be further used for recurrence analysis of fluctuations exhibited in the nonlinear distributions of the semi-variogram series.

### 3.2 Fuzzy recurrence plots

Stemming from the concept of recurrence plots (RPs) developed in chaos and nonlinear dynamics as a descriptive statistical technique for quantifying recurrences occuring in a trajectory [11], the method of fuzzy recurrence plots (FRPs) was introduced [12]. An FRP can be constructed from time series based on the concepts of artificial intelligence (AI) known as fuzzy logic [13] and its cluster analysis [14]. It has been shown that, in comparison with the method of FRs, an FRP can quantify recurrences and present visual of a system underlain with nonlinear behavior in a more natural way.

An FRP is mathematically described as follows. Let **x** = (*x*_1_, *x*_2_, …, *x*_*T*_) be a sequence of the semi-variograms of a mandibular CT scan, where *T* is the number of CT slices. Let *m* and *τ* be the embedding dimension and time delay for the construction of an FRP, respectively. The phase space of the series **x**, denoted as **Y**, is expressed as

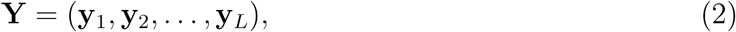

where *L* = *T* − (*m* − 1)*τ*, and its elements are formed by

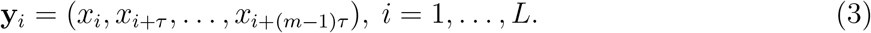

Using the phase space and fuzzy *c*-means (FCM) algorithm [14], an FRP is computed as follows. Given a number of clusters *c*, the FCM partitions **Y** into *c* clusters: (**v**_1_, **v**_2_, …, **v**_*c*_). The FCM also associates each element of **Y** with real membership grades in [0, 1], which expresses degrees of similarity between each element and all clusters, denoted as *µ*(**y**_*i*_, **v**_*k*_), *i* = 1, …, *L, k* = 1, …, *c*. Here, a larger membership value indicates a higher degree of the recurrence of the phase space. As the result, an FRP, denoted as **R**, is formulated as

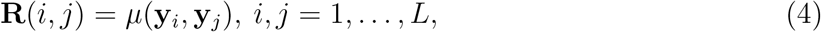

where *µ*(**y**_*i*_, **y**_*j*_) is the degree of similarity between two elements **y**_*i*_ and **y**_*j*_, which can be inferred based on the fuzzy similarity relations [16] as follows.

1. Reflexivity (self-similar): *µ*(**y**_*i*_, **y**_*i*_) = 1, *i* = 1, …, *L*.
2. Symmetry (computed by FCM): *µ*(**y**_*i*_, **v**_*k*_) = *µ*(**v**_*k*_, **y**_*i*_), *i* = 1, …, *L*; *k* = 1, …, *c*
3. Transitivity (inferred by fuzzy reasoning): *µ*(**y**_*i*_, **y**_*j*_) = max [min*{µ*(**y**_*i*_, **v**_*k*_), *µ*(**v**_*k*_, **y**_*j*_)*}*], *k* = 1, …, *c*; *i* ≠ *j*.

### 3.3 Fuzzy recurrence networks

Network theory in graph theoretic analysis has been applied to medicine known as network medicine, which is an emerging research area for studying human diseases [17] and processing big data in biomedicine [18]. Here, networks of fuzzy recurrence [15] is adopted to construct network topology and compute graph properties of the mandible on CT.

Using the same fuzzy similarity relations [16], the membership grades of recurrence or similarity between cluster pairs (**v**_*k*_, **v**_*q*_) can be obtained as

1. Reflexivity (self-similar): (**v**_*k*_, **v**_*k*_) = 1, *k* = 1, …, *c*.
2. Symmetry (computed by FCM): *µ*(**v**_*k*_, **y**_*i*_) = *µ*(**y**_*i*_, **v**_*k*_), *k* = 1, …, *c*; *i* = 1, …, *L*.
3. Transitivity (inferred by fuzzy reasoning): *µ*(**v**_*k*_, **v**_*q*_) = max [min{*µ*(**v**_*k*_, **y**_*i*_), *µ*(**y**_*i*_, **v**_*q*_)}], *i* = 1, …, *L*; *k* ≠ *q*.

A defuzzification procedure allows the transformation of an FRN into a binary network using the *α*-cut method by

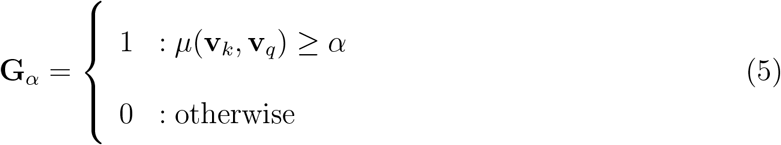

where **G**_*α*_ is a *c* × *c* binarized matrix, and *α* takes values in [0,1].

Finally, the adjacency matrix of an unweighted *α*-cut recurrence network of size *c* × *c*, denoted as **A**_*α*_, is defined as follows:

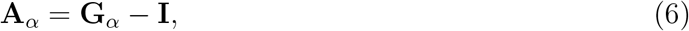

where **I** is the identity matrix of size *c* × *c*, and **A**_*α*_ can be used to compute graph properties such as the characteristic path length and average clustering coefficient [19, 20, 21].

### 3.4 Fuzzy recurrence quantification

An FRP **R**, who elements take real values in [0, 1], can be transformed into a binary FRP, denoted as **B**, using an image segmentation method described in [22]. Information provided by both **R** and **B** can be used to quantify recurrence of a dynamical system, which are described as follows [22].

#### 3.4.1 Fuzzy recurrence rate

Fuzzy recurrence rate, denoted as *f RR*, quantifies the rate of recurrences in **R** as

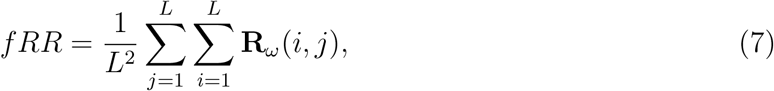

where **R**_*ω*_(*i, j*) = 0 for **R**(*i, j*) *< ω, i, j* = 1, …, *L*; implying recurrence points are considered for only FRP elements whose values *≥ ω*.

#### 3.4.2 Fuzzy determinism

Fuzzy determinism (*f DET*) quantifies recurrence with respect to lines along the main diagonal in **B**, which is expressed as

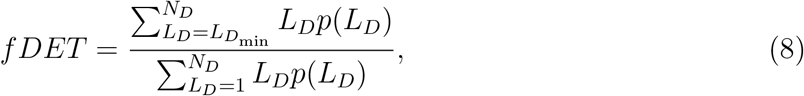

with *L*_*D*_ = diagonal length, 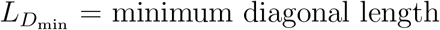, and *N*_*D*_ = maximum diagonal length of **B**; *N*_*D*_ = main diagonal length -1, and *p*(*L*_*D*_) is the probability of *L*_*D*_ computed from the histogram with *N*_*D*_ bins.

#### 3.4.3 Fuzzy laminarity

Fuzzy laminarity (*f LAM*) is the rate of recurrence with respect to vertical lines in **B**, which is defined as

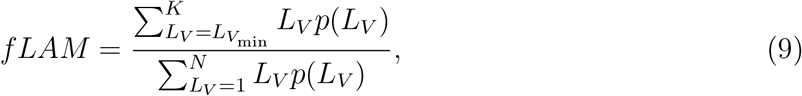

where *L*_*V*_ = diagonal length, 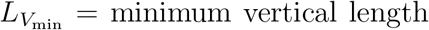, and *K* = maximum vertical length of **B**; and *p*(*L*_*V*_) is the probability of *L*_*V*_ obtained from the histogram with *K* bins.

#### 3.4.4 Fuzzy trapping time

Fuzzy trapping time (*f TT*) quantifies the average length of vertical lines in **B** as

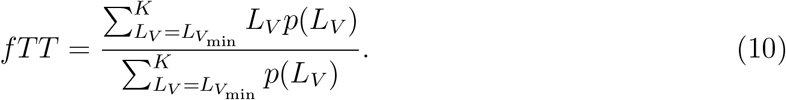

#### 3.4.5 Fuzzy divergence

Fuzzy divergence (*f DIV*) measures the divergence of the phase-space trajectory, which is expressed as

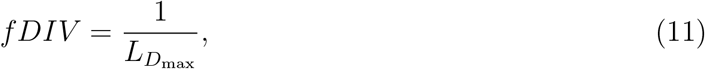

where 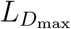 = maximum diagonal length of **B**.

#### 3.4.6 Fuzzy recurrence entropy

Fuzzy recurrence entropy (*f ENT*) is computed as the Shannon entropy of the diagonal lines in **B**:

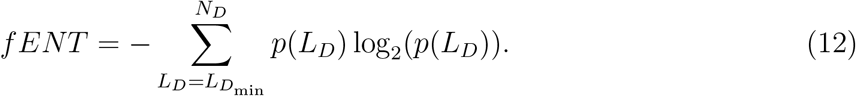

### 3.5 Largest recurrence eigenvalue

The determination of the largest recurrence eigenvalue [23], denoted as *λ*_max_, is described as follows:

1. Input **R** of size *L* × *L*.
2. Specify a filter kernel, ReLU, pool size, and stride
3. Specify *n* as final *n* × *n* convolved matrix *c***R**
4. While *L > n*
5. Convole **R** with ReLU
6. Do max pooling on the *c***R**
7. End While loop
8. Compute *λ*_max_ of the final *c***R**.

The ReLU, denoted as *u*, is defined as

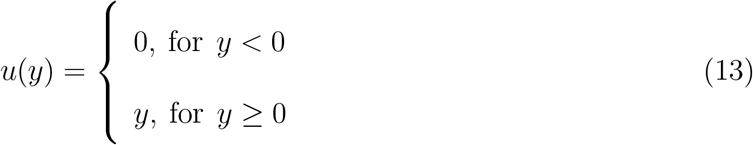

The maximum pooling works as follows. Given a pooling size *s* × *s*, and a set of pooling regions **P** = (**P**_1_, **P**_2_, …, **P**_*w*_, …, **P**_*W*_), where **P**_*w*_ = (*z*_*w*,1_, *z*_*w*,2_, …, *z*_*w,s*×*s*_). The maximum pooling, denoted as *H*_max_, that operates on a pooling size *s* × *s* is defined as

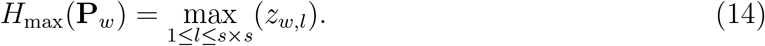

The number of pooling regions within a *c***R** is determined by the pool size and stride, which is the step size for traversing the *c***R**.

## 4 Results and Discussion

Figure 2 shows the plots of semi-variogram series obtained from the first 100 slices of the CT scans of the 10 subjects. The selection of this number of slices was due to the total number of slices otained from subject S3 = 110, where the last 10 slices contains little image information about the mandible. The semi-variogram values were calculated using *h* = 1 (to maximize the measure of spatial autocorrelation), and only voxel intensity *≥* 0.5 ∈ [0, 1] from row 150th were included in order to avoid a large part of the image background and non-mandible regions. Figure 2 shows the sequential spatial variances captured from the CT scans of the mandibles. The semi-variogram series of the male subjects tend to exhibit higher values than those of the female subjects, showing a pattern of dissimilarity in mandibles between genders.

**Figure 2:**
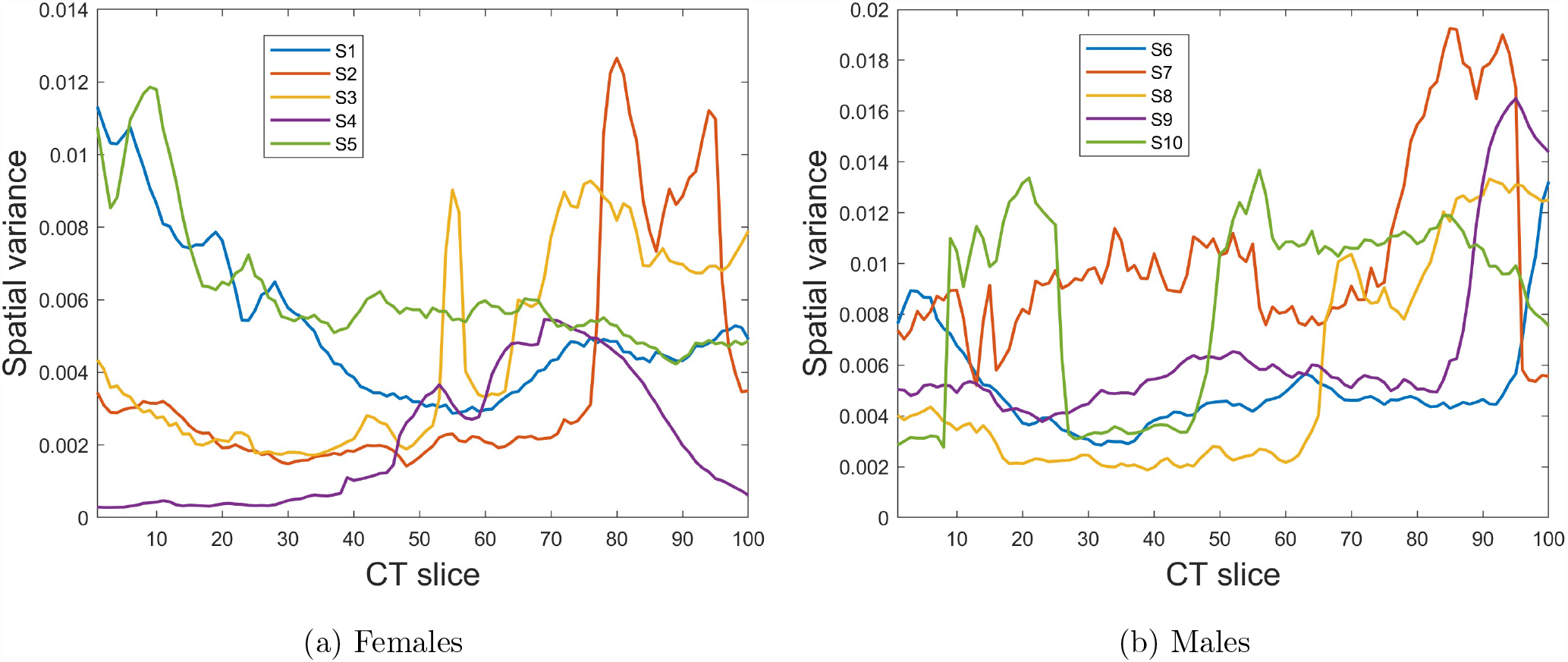
Semi-variograms of mandibles on CT.

Fluctuations of the semi-variogram sequences as depicted from Figure 2 lend themselves to nonlinear data analysis using the method of FRPs. Figure 3 shows the FRPs constructed from the 10 semi-variogram sequences shown in Figure 2. The FRPs were computed using embedding dimension *m* = 3, time delay *τ* = 1, and number of clusters *c* = 10.

**Figure 3:**
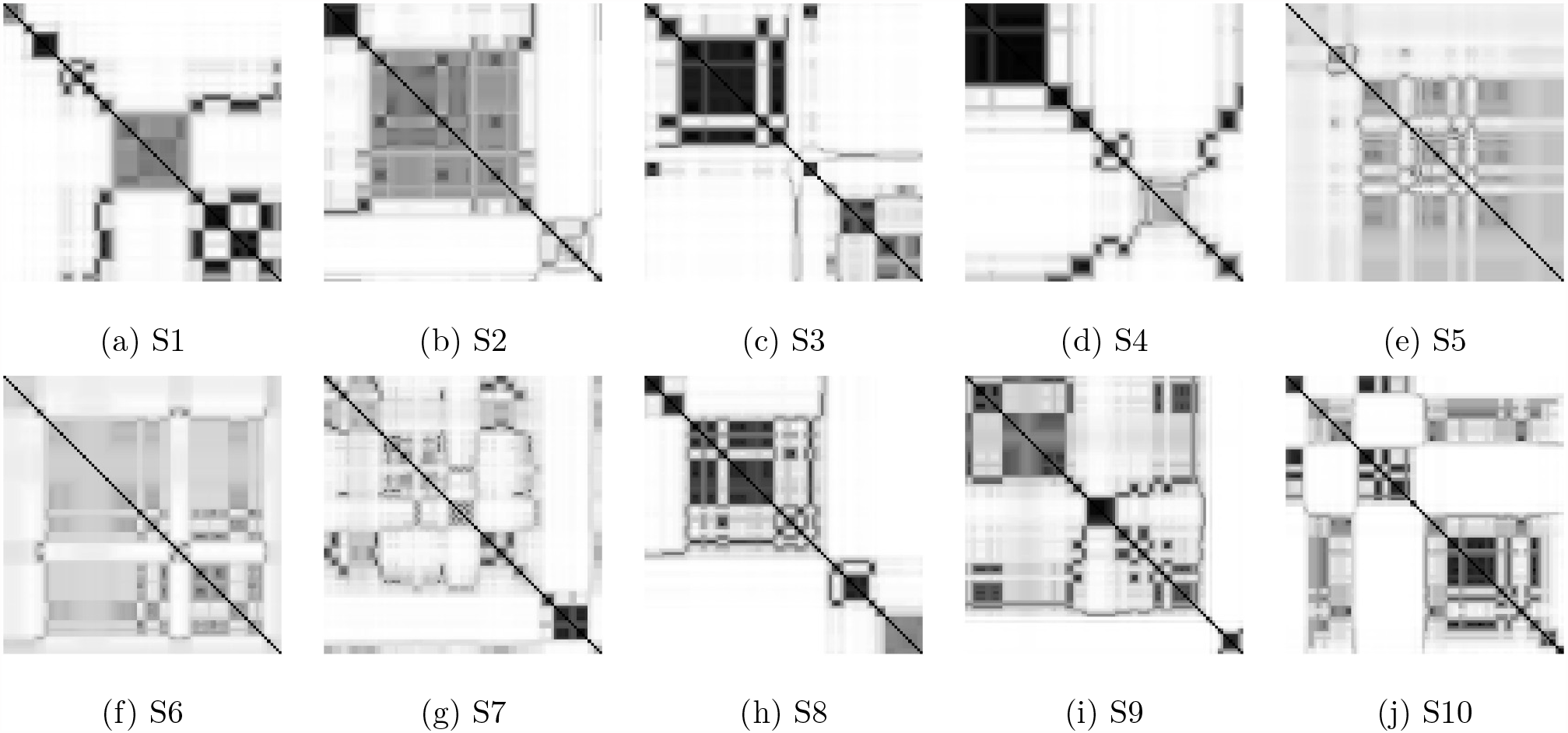
FRPs of mandibles on CT: females (first row), and males (second row).

In general, either RPs or FRPs are characterized with large-scale (typology) and small-scale (texture) patterns. The typology consists of homogeneous, periodic, drift, and disrupted/abrupt types. A homogeneous FRP indicates a stationary system that is governed by a random time series. An FRP that displays diagonal lines and checkerboards refers to a periodic time series, including a quasi-periodic signal whose FRP shows different distances between the diagonal lines. The typology of a drift, which is caused by a slowly varying dynamical system, displays changes in the upper-left and lower-right corners of an FRP. The disrupted typology, which shows white bands in an FRP, expresses abrupt changes in a dynamical system and can be useful for studying extreme and rare events. Here, the FRPs constructed from all CT scans exhibit abrupt changes of the spatial autocorrelation. However, all the FRPs are of different visuals. To quantify the recurrence patterns of the FRPs, several measures were calculated and shown in Table 2, where values for the minimum vertical and diagonal lines = 5. The mean values of *f RR, fTT*, and *f DIV* are different between male and female subjects.

**Table 2:**
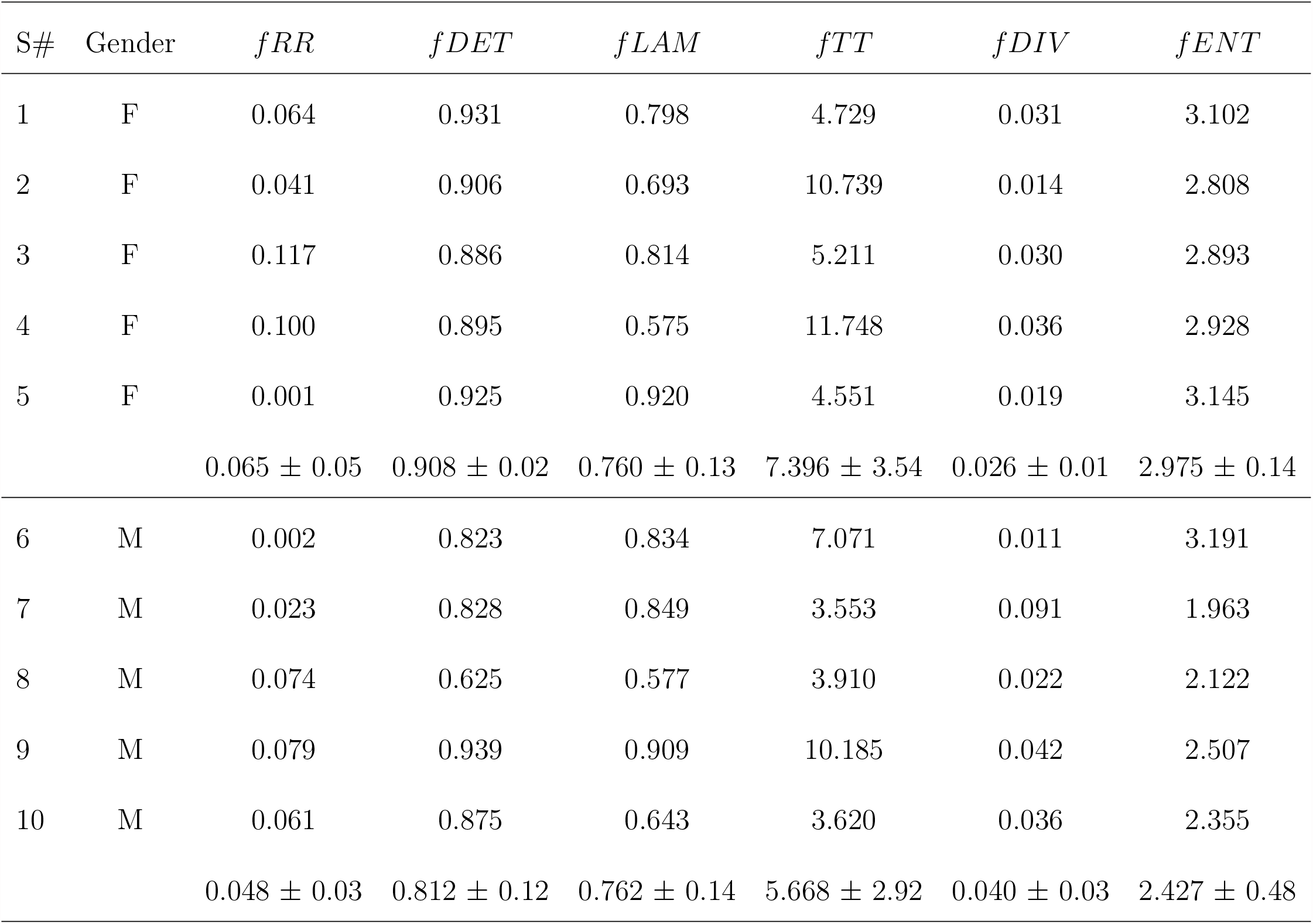
Recurrence quantification of mandibles on CT slices.

| S# | Gender | $fRR$ | $fDET$ | $fLAM$ | $fTT$ | $fDIV$ | $fENT$ |
| --- | --- | --- | --- | --- | --- | --- | --- |
| 1 | F | 0.064 | 0.931 | 0.798 | 4.729 | 0.031 | 3.102 |
| 2 | F | 0.041 | 0.906 | 0.693 | 10.739 | 0.014 | 2.808 |
| 3 | F | 0.117 | 0.886 | 0.814 | 5.211 | 0.030 | 2.893 |
| 4 | F | 0.100 | 0.895 | 0.575 | 11.748 | 0.036 | 2.928 |
| 5 | F | 0.001 | 0.925 | 0.920 | 4.551 | 0.019 | 3.145 |
| | | $0.065 \pm 0.05$ | $0.908 \pm 0.02$ | $0.760 \pm 0.13$ | $7.396 \pm 3.54$ | $0.026 \pm 0.01$ | $2.975 \pm 0.14$ |
| 6 | M | 0.002 | 0.823 | 0.834 | 7.071 | 0.011 | 3.191 |
| 7 | M | 0.023 | 0.828 | 0.849 | 3.553 | 0.091 | 1.963 |
| 8 | M | 0.074 | 0.625 | 0.577 | 3.910 | 0.022 | 2.122 |
| 9 | M | 0.079 | 0.939 | 0.909 | 10.185 | 0.042 | 2.507 |
| 10 | M | 0.061 | 0.875 | 0.643 | 3.620 | 0.036 | 2.355 |
| | | $0.048 \pm 0.03$ | $0.812 \pm 0.12$ | $0.762 \pm 0.14$ | $5.668 \pm 2.92$ | $0.040 \pm 0.03$ | $2.427 \pm 0.48$ |

To construct FRP-based networks for mandibles on the 10 CT scans, *α* = 0.4 and *β* = 0.08 were used. Figure 4 shows the network topologies generated from the FRPs of the CT scans. Each network topology is unique among each other. The characteristic path lengths and average clustering coefficients of the networks are shown in Table 3. There is a slight difference between the mean values of the characteristic path lengths for male (0.017 ± 0.02) and female (0.019 ± 0.03) subjects. The mean values of the average clustering coefficients for male (0.050 ± 0.01) and female (0.050 ± 0.02) subjects are similar.

**Table 3:**
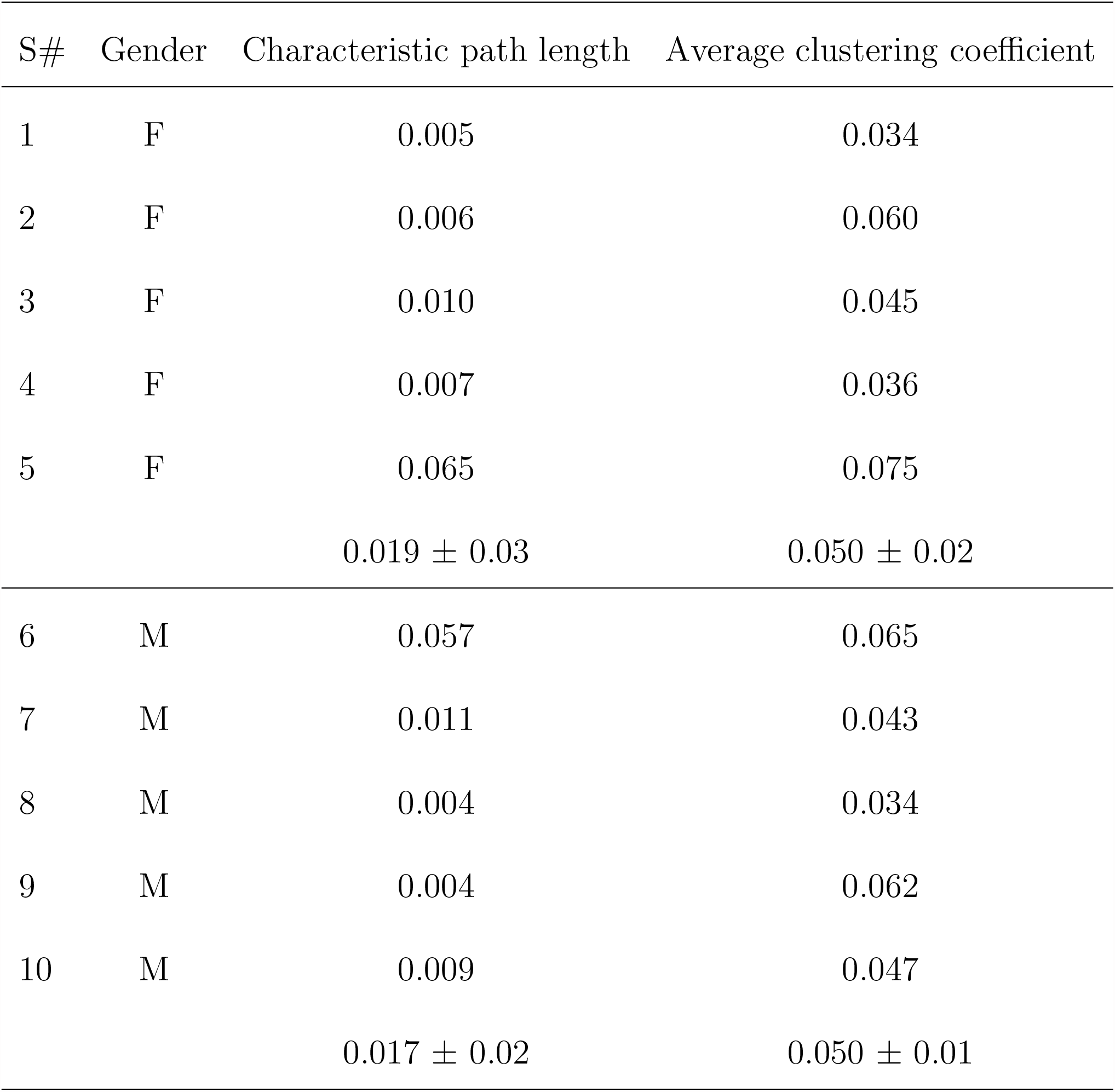
Graph properties of mandibles on CT slices.

**Figure 4:**
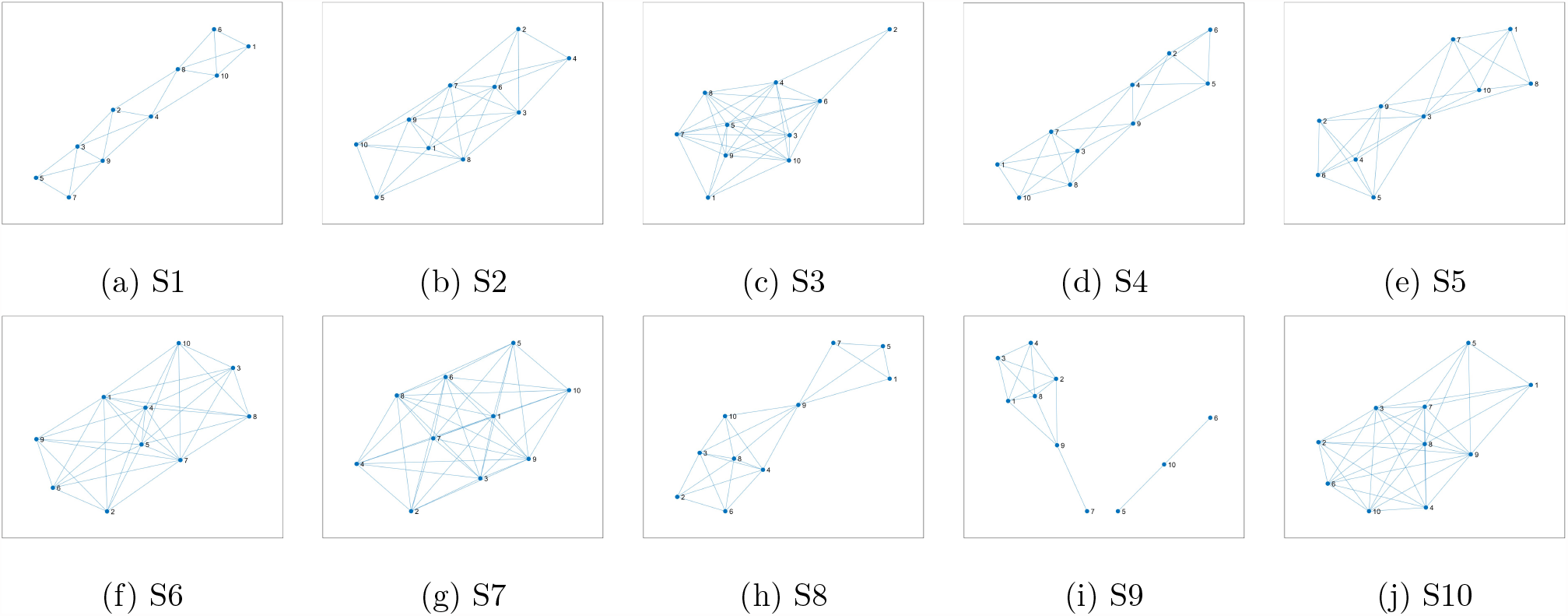
FRNs of mandibles on CT: females (first row), and males (second row).

Table 4 shows the largest recurrence eigenvalues of the FRPs of mandibles on 10 CT scans obtained from the 10 subjects. The largest recurrence eigenvalues were computed using the 3 × 3 convolutional kernel = [0 -1 0; -1 5 -1; 0 -1 0] (image sharpening), pooling size = 5, and final size of convolved matrix = 70. The largest eigenvalues for the male mandibles tend to be smaller than those for the female subjects. However, the mean largest eigenvalues between the two genders are quite similar (9.0 ± 2.2 for male and 9.2 ± 1.5 for female).

**Table 4:** Largest recurrence eigenvalues of mandibles on CT slices.

| S# | Gender | $\lambda_{\max}$ |
| --- | --- | --- |
| 1 | F | 7.952 |
| 2 | F | 11.270 |
| 3 | F | 8.053 |
| 4 | F | 10.095 |
| 5 | F | 8.450 |
| | | $9.164 \pm 1.50$ |
| 6 | M | 9.393 |
| 7 | M | 12.727 |
| 8 | M | 7.164 |
| 9 | M | 7.777 |
| 10 | M | 7.937 |
| | | $9.000 \pm 2.24$ |

The mandible has been utilized as an evidence for forensic investigations and archaeological studies in terms of sex identification when cranial and pelvic bones are absent [24]. Utilization of mandibular bones for the expedition of human identification was reported in the literature of oral and maxillofacial pathology [25], where the ramus of the mandible can be used for identifying gender and age in forensic science. Statistical analysis of morphological variation in the human mandible was carried out for classifying sex and ancestry in unknown subjects [26]. It was further reported that 10 mandibles had been used for gender classification using both linear and mixture discriminant methods [27]. The results showed that the analysis of mandible data would be useful to the communities of anthropology and forensics. In particular, the ability for determining sex by means of the mandible plays an important role in forensics. Thus, machine-leaning methods have been applied for sex identification using volumetric and linear measurements of the mandible on cone beam CT scans [28]. Dental biometrics involves the utilization of dental radiographs for human identification, emerging as a cutting-edge technology in forensic data analysis, where other human remains such as faces, fingerprints, and irises are missing due to injuries caused by accidents or natural disasters. Mandibular information was extracted from panoramic dental radiographs and outer contours of mandibles were used as time series of spatial mandibular structures for matching and recognizing individuals using an autoregressive model [29]. In line with previous studies, the work reported herein can contribute to the aspect of image feature extraction for machine learning to perform computer-based identification of gender and individual identity.

Furthermore, the approach proposed in this study can be applied for discriminating subtle differences between individual faces. Such a technical capability is essential for social understanding by solving the general challenge of facial recognition. A study has recently been reported to use 3D morphable models based on principal component analyses of real faces [30]. The study was able to discern differences between faces and to generate new faces. Features of the mandible on CT scans developed in this study can be used with deep learning models, which are a state-of-the-art AI approach, for facial recognition and generation.

## 5 Conclusion

Methods of AI-based nonlinear dynamics and network analysis for extracting spatial information of the mandible on CT have been presented in the foregoing sections. As a proof of concept, results obtained from testing the proposed approach with 10 CT scans from 5 male and 5 female subjects suggested potential features of dissimilarity and similarity between male and female subjects. The finding highlights unique characteristics of human mandibles captured on radiographic data. Due to the small data size, this study was unable to investigate associations among mandibles, ages, and body weights. More data are needed to rigorously confirm the present finding and allow investigations of the mentioned relationships of the mandible with other physical variables.

Furthermore, it is of clinical interest to apply the proposed approach for exploring complex image properties of human mandibles for improving the diagnostics and treatment of facial trauma. It has recently been reported that by considering the uniqueness of the mandible can enhance clinical approaches to mandibular regeneration in order to avoid postsurgical complications [1]. In parallel to precision medicine [31], precision dentistry [32, 33] is an emerging area of research that has potential to achieve precise dental diagnostics and targeted treatment. Important roles of AI and image analysis for contribution to precision dentistry have been quickly realized [34]. The feature extraction and network analysis proposed in this study can be relevant to gaining insights into dentomaxillofacial imaging.

## Data Availability

All data produced are available online at Figshare
https://doi.org/10.6084/m9.figshare.6167726.v5

## Ethics

This work did not require ethical approval from a human subject or animal welfare committee.

## Data and code accessibility

This article has no additional data. The Matlab codes implemented in this study are available at the first author’s personal website https://sites.google.com/view/tuan-d-pham/codes under the title “Features and Networks of the Mandible”.

## Authors’ contributions

TDP conceptualized the work, developed methods of image and network analysis, and wrote the paper. SBH, MP, and PC contributed clinical knowledge, result analysis, and editing. All authors read and approved the submitted version.

## References

[1] Soares AP, Fischer H, Aydin S, Steffen C, Schmidt-Bleek K, Rendenbach C. Uncovering the unique characteristics of the mandible to improve clinical approaches to mandibular regeneration. Front Physiol. 14, 1152301, 2023.

[2] Vinay G, Gowri MSR, Anbalagan J. Sex determination of human mandible using metrical parameters. J Clin Diagn Res. 7, 2671–2673, 2013.

[3] Fan Y, Penington A, Kilpatrick N, Hardiman R, Schneider P, et al. Quantification of mandibular sexual dimorphism during adolescence. J Anat. 234, 709–717, 2019.

[4] Hazari P, Hazari RS, Mishra SK, Agrawal S, Yadav M. Is there enough evidence so that mandible can be used as a tool for sex dimorphism? A systematic review. J Forensic Dent Sci. 8, 174, 2016.

[5] Chole RH, Patil RN, Chole SB, Gondivkar S, et al. Association of mandible anatomy with age, gender, and dental status: A radiographic study. International Scholarly Research Notices 2013, article 453763, 2013.

[6] Maresky HS, Klar MM, Tepper J, Gavriel H, Ziv Baran T, et al. Mandibular width as a novel anthropometric measure for assessing obstructive sleep apnea risk. Medicine (Baltimore) 98, e14040, 2019.

[7] Cerda C, Lezcano MF, Marinelli F, Alarcon J, Fuentes R. Determination of mandibular position and mouth opening in healthy patients and patients with articular and/or muscular pathology: A pilot study with 3D electromagnetic articulography and surface electromyography. Journal of Clinical Medicine 12, 4822, 2023.

[8] Olayemi AB. Assessment and determination of human mandibular and dental arch profiles in subjects with lower third molar impaction in Port Harcourt, Nigeria. Ann Maxillofac Surg. 1, 126–130, 2011.

[9] Wallner J, Mischak I, Egger J. Computed tomography data collection of the complete human mandible and valid clinical ground truth models. Sci. Data 6, 190003, 2019.

[10] Wallner J, Egger J. Mandibular CT dataset collection. Figshare, https://doi.org/10.6084/m9.figshare.6167726.v5, 2018.

[11] Eckmann JP, Kamphorst SO, Ruelle D. Recurrence plots of dynamical systems. Europhysics Letters 5, 973–977, 1987.

[12] Pham TD. Fuzzy recurrence plots. EPL 116, 50008, 20116.

[13] Zadeh LA. Fuzzy sets. Information and Control 8, 338–353, 1965.

[14] Bezdek JC, Ehrlich R, Full W. FCM: The fuzzy c-means clustering algorithm. Computers & Geosciences 10, 191–203, 1984.

[15] Pham TD. From fuzzy recurrence plots to scalable recurrence networks of time series. EPL 118, 20003, 2017.

[16] L.A. Zadeh. Similarity relations of fuzzy orderings. Inform. Sci., 3 (1971), p. 177–200.

[17] Barabasi AL, Gulbahce N, Loscalzo J. Network medicine: a network-based approach to human disease. Nat Rev Genet 12, 56–68, 2011.

[18] Sonawane AR, Weiss ST, Glass K, Sharma A. Network medicine in the age of biomedical big data. Front Genet. 10, 294, 2019.

[19] Watts DJ, Strogatz S. Collective dynamics of “small-world” networks. Nature 393, 440–442, 1998

[20] Albert R, Barabasi AL. Statistical mechanics of complex networks. Rev. Mod. Phys. 74, 47–97, 2002.

[21] Pham TD, Fan C, Pfeifer D, Zhang H, Sun XF. Image-based network analysis of DNp73 expression by immunohistochemistry in rectal cancer patients. Frontiers in Physiology 10, 1551, 2019.

[22] Pham TD. Quantification analysis of fuzzy recurrence plots. EPL 137, 62002, 2022.

[23] Pham TD. Convolutional fuzzy recurrence eigenvalues. EPL 135, 20002, 2021.

[24] Varela LM, Moss BH, Moore-Jansen P. Morphological variation in the mandible of white males and females from the East Texas region for potential applications for skeletal identification. Canadian Society of Forensic Science Journal 55, 181–205, 2022.

[25] Singh S, Bavle RM, Konda P, Venugopal R, Bopaiah S, Kumar S. Assessment of the most reliable sites in mandibular bone for the best deoxyribonucleic acid yield for expeditive human identification in forensics. J Oral Maxillofac Pathol. 21, 447–453, 2017.

[26] Berg GE. Biological Affinity and Sex Determination using Morphometric and Morphoscopic Variables from the Human Mandible. PhD diss., University of Tennessee, TN: Knoxville, 2008.

[27] Berg GE, Kenyhercz MW. Introducing human mandible identification [(hu)MANid]: A free, web-based GUI to classify human mandibles. J Forensic Sci 62, 1592–1598, 2017.

[28] Baban MTA, Mohammad DN. The accuracy of sex identification using CBCT morphometric measurements of the mandible, with different machine-learning algorithms–A retrospective study. Diagnostics 13, 2342, 2023.

[29] Banday M, Mir AH. Dental biometric identification system using AR model, 2019 IEEE Region 10 Conference (TENCON 2019), Kochi, India, 2019, pp. 2363–2369.

[30] Jozwik KM, O’Keeffe J, Storrs KR, Guo W, Golan T, Kriegeskorte N. Face dissimilarity judgments are predicted by representational distance in morphable and image-computable models. Proc Natl Acad Sci U S A 119, e2115047119, 2022.

[31] Ginsburg GS, Phillips KA. Precision medicine: From science To value. Health Aff (Millwood) 37, 694–701, 2018.

[32] Schwendicke F, Krois J. Precision dentistry–what it is, where it fails (yet), and how to get there. Clin Oral Invest 26, 3395–3403, 2022.

[33] Kaur N, Mishra G, Parihar V, Sharma SS, Tyro KD, Mann SK. Precision dentistry. Br Dent J 234, 197, 2013.

[34] Hung KF, Yeung AWK, Bornstein MM, Schwendicke F. Personalized dental medicine, artificial intelligence, and their relevance for dentomaxillofacial imaging. Dentomaxillofac Radiol 52, 20220335, 2023

